# Association between pregnancy and severe COVID-19 symptoms in Qatar: a cross-sectional study

**DOI:** 10.1101/2022.07.20.22277847

**Authors:** Alla’ K. Al-Qassem, Ammar B. Humaidi, Amna K. Al-Kuwari, Elham M. Hasan, Nosaiba H. Yakti, Rakan M. Al-Hathal, Devendra Bansal, Elmoubashar Abu Baker Abd Farag, Hamad E. Al-Romaihi, Mohammed H. J. Al-Thani, Omran A. H. Musa, Suhail Doi, Tawanda Chivese

## Abstract

**Background:** There is inconclusive evidence whether pregnancy exacerbates COVID-19 symptoms or not, and scarce data from the Middle East and North Africa region. The aim of this study was to investigate the association between pregnancy and COVID-19 symptoms in Qatar.

**Methods:** This cross-sectional study was carried out using data of all women with confirmed COVID-19, comparing pregnant and non-pregnant women of child-bearing age (18-49 years). Data of all COVID-19 cases were collected by the Ministry of Public Health (MoPH) in Qatar, between March and September 2020. Symptoms were compared by pregnancy status and classified into moderate and severe. Multivariable logistic and poisson regression was carried out to investigate the association between pregnancy and severity of COVID-19 symptoms.

**Results:** During the study period, 105744 individuals were diagnosed with COVID-19, 16908 were women of childbearing age. From that sample, 799 women who were pregnant (mean age 29.9 years (SD 5.2)) and 16109 women who were not pregnant (mean age 33.1 years (SD 7.8)). After multivariable logistic regression, pregnancy was associated with a 1.4-fold higher odds of reporting any symptoms of COVID-19 (OR 1.41, 95% CI 1.18-1.68), and 1.3-fold higher odds of reporting shortness of breath (OR 1.29, 95% CI 1.02-1.63). After multivariable poisson regression, pregnancy was also associated with a higher number of symptoms (IRR 1.03, 95%CI 0.98-1.08).

**Conclusion:** Our findings suggest that, in this setting, pregnant women are more likely to have symptomatic COVID-19, and shortness of breath, compared to non-pregnant women of childbearing age.

## Introduction

Pregnant women may be at greater risk of infectious diseases, due to the physiological and immunological changes which occur during pregnancy, that can impair pathogen clearance and consequently increase disease severity [1]. Based on this, there are reasonable concerns that COVID-19 could be severe during pregnancy and possibly cause adverse pregnancy outcomes [2]. A severe course of COVID-19 may result in adverse pregnancy outcomes such as stillbirths and maternal deaths [3]. It may therefore be necessary to prioritize protections for pregnant women if they are at higher risk of severe COVID-19.

Although mounting evidence suggests that pregnant women are more susceptible to severe COVID-19, findings on the association between pregnancy and the severity of COVID-19 are not conclusive. One systematic review and a meta-analysis found that pregnant women were at a greater risk of requiring ventilatory support or becoming admitted to an intensive care unit (ICU), in comparison with non-pregnant women of childbearing age [4]. Another meta-analysis showed an increased risk of severe COVID-19 and a worse symptom profile in pregnant women [5]. In contrast, one systematic review found no association between COVID-19 severity, assessed using symptoms and pregnancy. In this review, most pregnant women had mild symptoms, and the clinical presentation of pregnant women was similar to that of the general population [6]. Similarly, another systematic review also reported that certain symptoms such as cough, fatigue, sore throat, headache, and diarrhea to be less probable in pregnant than in non-pregnant women [7]. These findings suggest the need for more research on this topic.

Even though COVID-19 is predicted to become endemic, and to have a more benign course due to the high levels of population immunity, availability of vaccines and prior infection [8], there are still individuals who are at risk of severe symptomatic COVID-19 [9]. Certain symptoms such as shortness of breath have been shown to be prognostic for severe COVID-19. For instance, one study showed that shortness of breath was associated with 2.4-folds odds of severe COVID-19 [10]. Other symptoms have also been shown to be associated with severe COVID-19 although the data are inconclusive [11]. Further, although it is likely that a higher symptoms count may imply a worse disease profile, there is little research on this.

This study investigated the association between pregnancy and COVID-19 symptoms in a Middle Eastern population where such data have been scarce to date. The primary objective of this study was to compare COVID-19 symptomatic status between women without pregnancy and pregnant women. We also compared individual symptoms between pregnant and non-pregnant women, and within age groups.

## Materials and methods

### Design and setting

This study was a cross sectional study of women of childbearing age with confirmed COVID-19 using data collected by the Ministry of Public Health (MoPH) in Qatar during the period of March to September 2020. Symptomatic status was evaluated across two groups, pregnant women, and women with no pregnancy. Additionally, in both groups individual symptoms were compared and categorized into mild to moderate and severe classes of symptoms according to a modified version of the criteria proposed by the National Institutes of Health (NIH) of the United States [12]. The study is reported following the Strengthening the Reporting of Observational Studies in Epidemiology (STROBE) (Supporting Information 1).

### Study participants

The population of Qatar consists of a large heterogeneous multinational population of which expatriates form the majority, around 90%, and the remainder are local [13]. This population was also reflected in the confirmed cases of COVID-19 from the MoPH. The MoPH collects data on all confirmed cases of COVID-19, diagnosed by polymerase chain reaction (PCR), in Qatar, including women of childbearing age both with and without pregnancy. For this study, participants were eligible if they were women aged between 18 to 49 years of age and if they had confirmed COVID-19 by RT-PCR. The study excluded individuals below 18 years or above 49 years of age. In addition, women in the postpartum period, defined as within 6 weeks after birth, were excluded.

### Data collection

From the MoPH dataset, we selected data for each participant which included symptoms of COVID-19, pregnancy status, sociodemographic data such as age, nationality, marital status, month of diagnosis, number of symptoms and comorbidities. The data on symptoms were collected by telephonic interview by the MoPH. Each individual was asked about presence of respiratory symptoms, such as cough, sore throat, and shortness of breath, gastrointestinal symptoms, such as diarrhea and abdominal pain, musculoskeletal symptoms that included muscle or joint pain, and systemic symptoms which consisted of fever, headache, and chills. The symptoms were classified into three categories. Category I, asymptomatic state, included those who had no symptoms. Category II, mild to moderate, comprised of participants who had fever, cough, sore throat, headache, muscle or joint pain, diarrhea, chills, or abdominal pain, but not shortness of breath. Category III, severe symptoms, consisted of participants who had shortness of breath. The severity categories were then compared between women without pregnancy and with pregnancy. We also compared individual symptoms by pregnancy status, and, further, compared both severity categories and individual symptoms by pregnancy status but within the age groups of 18-29 years, 30-39 years, and 40-49 years of age. Further, we categorized individual symptoms into organ systems and compared frequency across the three age groups.

### Statistical analysis

The categorical data in the study were summarized as frequencies and percentages, whereas age was normally distributed and therefore summarized using mean and standard deviation (SD). To compare the categorical data by pregnancy status, the chi-squared (χ2) test was utilized, and t-test was used to compare mean age. We utilized literature and directed acyclic graphs (Supplementary Fiigure 1) to identify potential confounders to adjust for the association between pregnancy and COVID-19, and in this case, only age appeared to meet the confounding criteria [14,15,16]. We also adjusted for diabetes, cardiovascular disease (CVD), and region of origin as these have been shown to be prognostic for severe COVID-19 [17,18]. We compared individual symptoms and symptom categories using the chi-squared test. Multivariable logistic regression was then carried out to assess the association between pregnancy and the severity of COVID-19 symptoms, adjusted for age. We carried out two models, the first model with the outcome being any symptomatic COVID-19 compared to no symptoms, second model with the outcome being shortness of breath compared to mild or no symptoms. The odds ratios (OR), 95% confidence intervals (95% CI), and exact p-values were reported. Additionally, we caried out multivariable poisson regression with the outcome being the number of symptoms and the exposure being pregnancy and adjusted for the same confounders. All analyses were carried out in Stata version 16 [19].

### Ethics approval

Ethics approval and waiver of informed consent were approved by the MoPH (ERC-826-3-2020), and all data were de-identified before the analysis.

## Results

### Characteristics of included participants

A total of 105 744 individuals were diagnosed with COVID-19 during the study period. From these, 17 610 were women of childbearing age. After exclusion of women in post-partum and those who were aged less than 18 years, 799 pregnant and 16 109 non-pregnant women were included (Fig 1). The characteristics of the included participants, by pregnancy status, are shown in Table 1.

**Table 1.** Characteristics of participants by pregnancy status.

| Variable |  | Overall,<br>n= 16908 | Pregnant,<br>n=799 | No<br>pregnancy,<br>n=16109 | P-value |
| --- | --- | --- | --- | --- | --- |
| Age, years | Mean (SD) | 33.0 (7.7) | 29.9 (5.2) | 33.1 (7.8) | <0.001 |
| Age groups, n (%) |  |  |  |  | <0.001 |
|  | 18-29 years | 5929<br>(35.1) | 380 (6.4) | 5549 (93.6) |  |
|  | 30-39 years | 7224<br>(42.7) | 393 (5.4) | 6831 (94.6) |  |
|  | 40-49 years | 3755<br>(22.2) | 26 (0.7) | 3729 (99.3) |  |
| Region of origin, n<br>(%) |  |  |  |  | <0.001 |
|  | Asia | 8105<br>(47.9) | 290 (3.6) | 7815 (96.4) |  |
|  | Middle east & north<br>Africa | 7746<br>(45.8) | 477 (6.2) | 7269 (93.8) |  |
|  | Sub-Saharan Africa | 767 (4.5) | 22 (2.9) | 745 (97.1) |  |
|  | North America &<br>Europe | 255 (1.5) | 8 (3.1) | 247 (96.9) |  |
|  | Australasia | 13 (0.1) | 0 (0.0) | 13 (100) |  |
|  | South America | 21 (0.1) | 2 (9.5) | 19 (90.5) |  |
| Marital status, n (%) |  |  |  |  | <0.001 |
|  | Married | 5917<br>(35.0) | 487 (8.2) | 5427 (91.8) |  |
|  | Single | 1763<br>(10.4) | 5 (0.3) | 1758 (99.7) |  |
|  | Divorced/widowed | 77 (0.5) | 0 (0.0) | 9 (100) |  |
|  | Not applicable | 9144<br>(54.1) | 295<br>(38.6) | 2069 (49.3) |  |
|  | Others | 9 (0.1) | 0 (0.00) | 9 (100) |  |
| Month of diagnosis,<br>n (%) |  |  |  |  | <0.001 |
|  | April | 546 (3.2) | 35 (6.4) | 511 (93.6) |  |
|  | May | 2816<br>(16.7) | 174 (6.2) | 2642 (93.8) |  |
|  | June | 4780<br>(28.3) | 202 (4.2) | 4578 (95.8) |  |
|  | July | 2468<br>(14.6) | 128 (5.2) | 2340 (94.8) |  |
|  | August | 3244<br>(19.2) | 139 (4.3) | 3105 (95.7) |  |
|  | September | 3054<br>(18.1) | 121 (4.0) | 2933 (96.0) |  |
| Number of<br>symptoms | Medium (IQR) | 2 (0-3) | 2 (0-3) | 2 (0-3) | 0.0138 |
| Death, n (%) |  | 0 (0.0) | 0 (0.0) | 0 (0.0) |  |

\*Values are numbers (percentages) unless stated otherwise.

**Figure 1.**
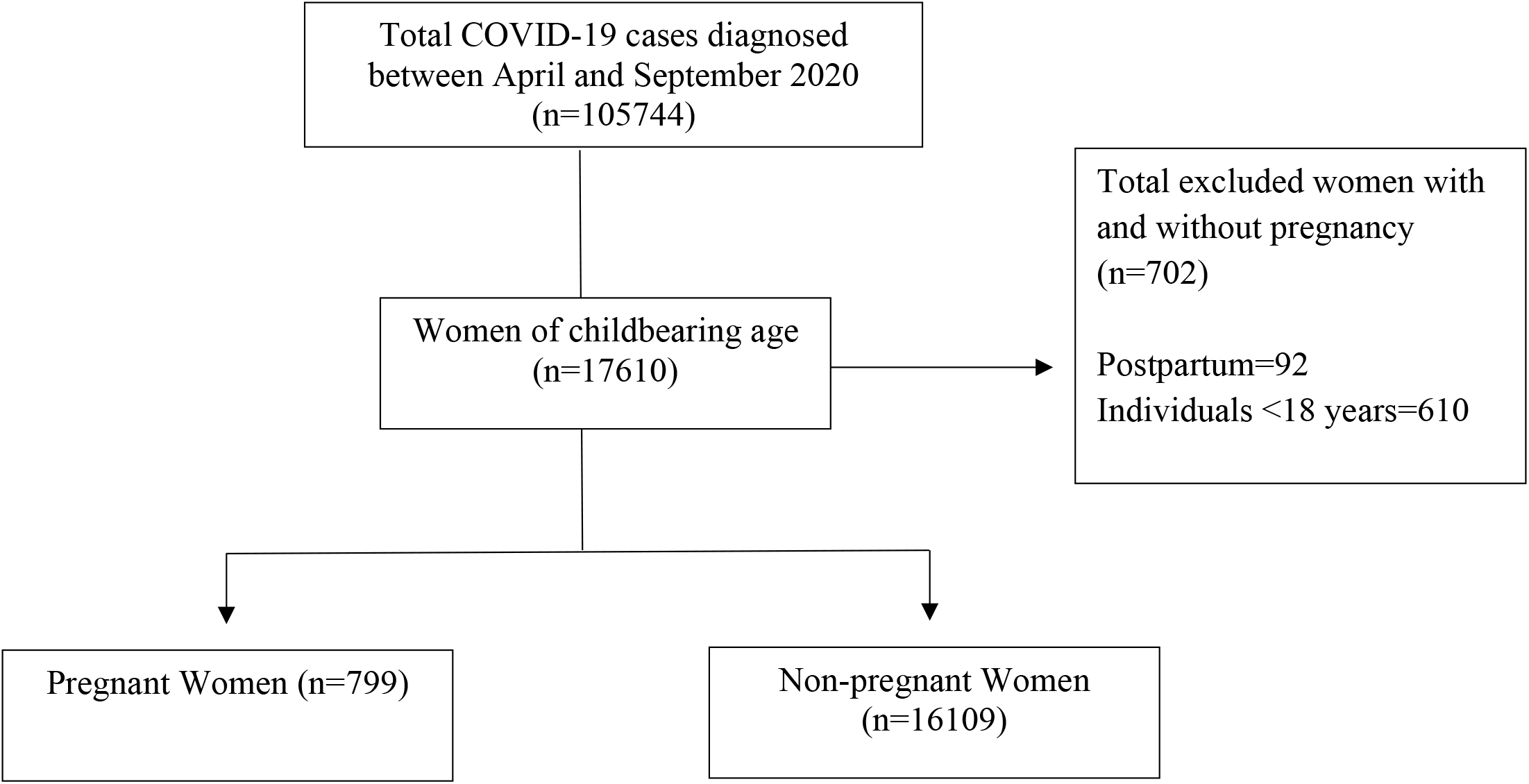
Study flow chart.

The mean age of the included women was 33.0 years (SD 7.7), with most of the individuals being aged between 30-39 years (42.7%) and 18-29 years (35.1%) (Table 1). Pregnant women were slightly younger than those without pregnancy (mean 29.9 years (SD 5.2) vs 33.1 years (SD 7.8), respectively, p<0.01). Most of the women were from countries in the Asia (47.9%), followed by countries in Middle East & North Africa (45.8%). Most of the participants were diagnosed in the months of June and August (28.3% and 19.2%, respectively). There were no deaths in both groups of women of childbearing age.

### Comparison of symptoms between pregnant and non-pregnant women

Table 2 compares COVID-19 symptoms between pregnant women and women with no pregnancy. Overall, the proportion of symptomatic COVID-19 was higher in the pregnancy group compared to the no pregnancy group (78.3% vs 70.5%, respectively, p<0.001), with strong evidence against the model hypothesis. In terms of individual symptoms, compared to women without pregnancy, a significantly higher proportion of pregnant women reported shortness of breath (10.6% vs 7.9%, respectively, p=0.006) and cough (37.3% vs 32.9%, p=0.016). No significant differences were observed between women with and without pregnancy across the other symptoms.

**Table 2.** Comparison of severity of symptoms between by pregnancy status.

|  |  | Pregnant women,<br>n=799 | No pregnancy,<br>n=16109 | P value |
| --- | --- | --- | --- | --- |
| Asymptomatic<br>n= 4920 | n(%) | 173(21.7) | 4747(29.5) | <0.001 |
| Symptomatic<br>n= 11988 | n(%) | 626(78.3) | 11362(70.5) | <0.001 |
| Mild to moderate group n= 10628 |  |  |  |  |
| Symptoms | Fever<br>n(%) | 301(42.2) | 6078(41.0) | 0.53 |
|  | Cough<br>n(%) | 266(37.3) | 4882(32.9) | 0.016 |
|  | Sore<br>throat<br>n(%) | 171(23.9) | 3587(24.2) | 0.89 |
|  | Headache<br>n(%) | 234(32.8) | 4644(31.3) | 0.41 |
|  | Muscle or<br>joint pain<br>n(%) | 211(29.6) | 3905(26.3) | 0.056 |
|  | Diarrhoea<br>n(%) | 31(4.3) | 855(5.8) | 0.11 |
|  | Chills<br>n(%) | 57(8.0) | 1069(7.2) | 0.43 |
|  | Abdominal<br>pain<br>n(%) | 49(6.9) | 842(5.7) | 0.18 |
| Severe group n=1360 |  |  |  |  |
| Severe symptoms | Shortness<br>of breath<br>n(%) | 85(10.6) | 1275(7.9) | 0.006 |

### Comparison of symptoms by pregnancy status within age groups

Table 3 shows a comparison of COVID-19 symptoms by pregnancy across different age categories. Overall, the proportion of symptomatic COVID-19 was significantly higher in pregnant compared to non-pregnant women in the age groups of 18-29 years (76.5% vs 69.6%, respectively, p<0.001), and 30-39 years (78.1% vs 70.8%, respectively, p=0.002). Shortness of breath was also reported by a higher proportion of pregnant women compared to women with no pregnancy in the age group of 18-29 years (13.2% vs 7.4%, respectively, p<0.001). With regards to mild to moderate symptoms, most symptoms were similar across both groups in the majority of the age categories, with few exceptions. These included cough (38.2% vs 32.7%, respectively, p<0.040) and muscle or joint pain (29.7% vs 24.3%, respectively, p=0.029) which were greater in pregnant women compared to women with no pregnancy in the age group of 18-29 years, respectively. Additionally, sore throat was more common in pregnant women than women without pregnancy in the age group 30-39 years (28.3% vs 23.3%, respectively, p=0.030). However, in the age group of 18-29 years, sore throat was of greater proportion in women without pregnancy compared to pregnant women in the age group of 18-29 years (25.7% vs 19.1%, respectively, p=0.008), Additionally, fever was more frequent in women with no pregnancy than it was in pregnant women in the age group of 40-49 years (41.4% vs 17.4%, respectively, p=0.020).

**Table 3.**
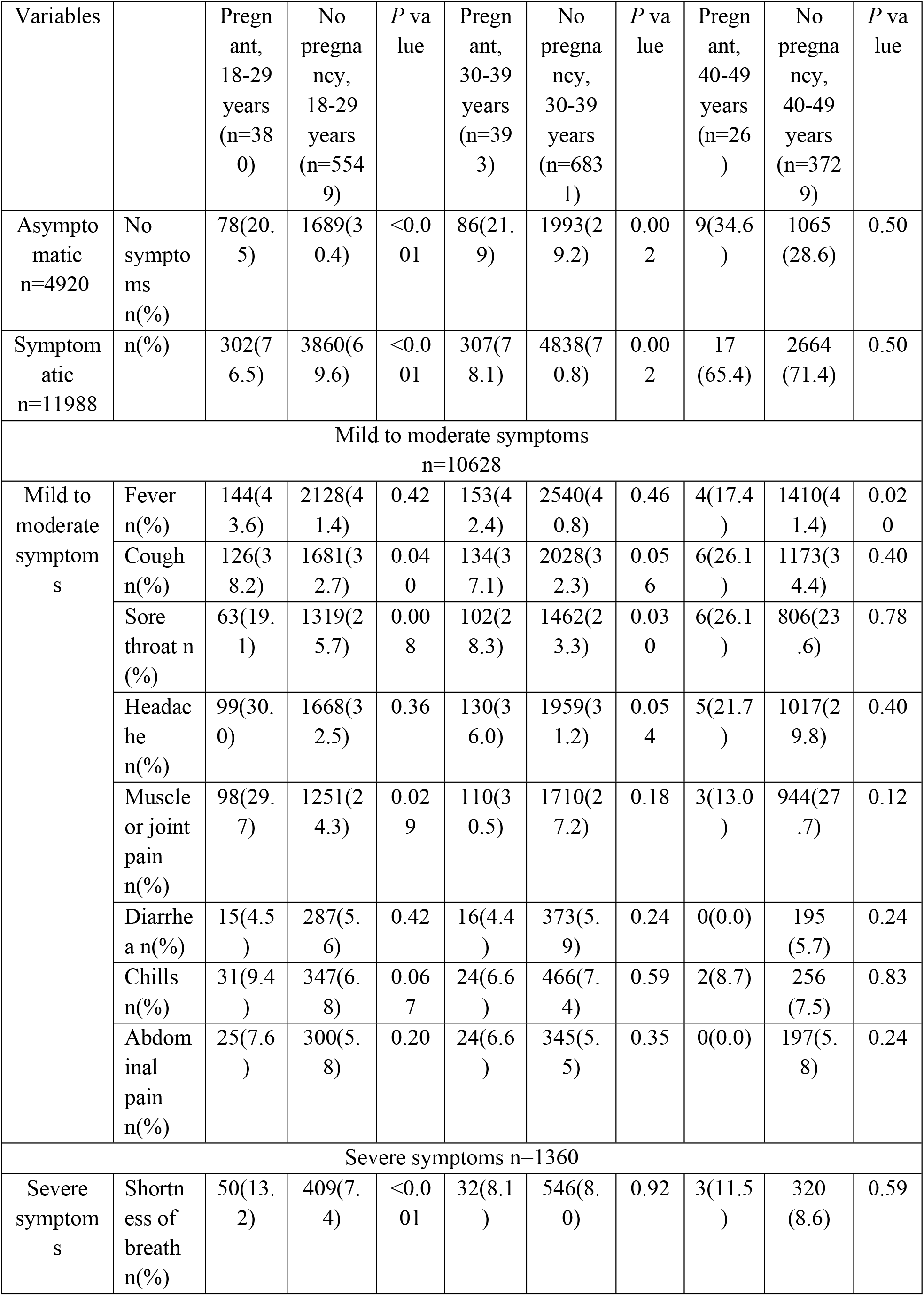
Comparison of severity of symptoms by pregnancy status in each age group.

### Comparison of symptoms categorized into organ systems between women with and without pregnancy

The COVID-19 symptoms that were reported by the three age groups of both cases and controls were further combined and categorized based on the organ system (Table 4). In general, respiratory symptoms were significantly higher in pregnant versus nonpregnant women in the age group of 18-29 years (52.9% vs 47.7%, respectively, p=0.050), and 30-39 years (52.9% vs 47.1%, respectively, p=0.023). In addition, musculoskeletal symptoms were more common in pregnant women than women with no pregnancy in age group 18-29 years (33.2% vs 26.8%, respectively, p=0.007). Other organ systems-based symptoms were fairly similar across both groups in most of the age categories.

**Table 4.** Comparison of organ systems-based symptoms by pregnancy status in each age group.

| Variables | Pregnant<br>18-29<br>years<br>(n%) | No<br>pregnancy<br>18-29<br>years<br>(n%) | P<br>value | Pregnant<br>30-39<br>years<br>(n%) | No<br>pregnancy<br>30-39<br>years<br>(n%) | P<br>value | Pregnant<br>40-49<br>years<br>(n%) | No<br>pregnancy<br>40-49<br>years<br>(n%) | P<br>value |
| --- | --- | --- | --- | --- | --- | --- | --- | --- | --- |
| Respiratory<br>symptoms<br>n% | 201(52.9) | 2647(47.7) | 0.050 | 208(52.9) | 3214(47.1) | 0.023 | 12(46.15) | 1824(48.9) | 0.779 |
| Gastrointestinal<br>symptoms<br>n% | 48(12.6) | 613(11.1) | 0.342 | 42(10.7) | 740(10.8) | 0.928 | 2(7.69) | 393(10.54) | 0.637 |
| Musculoskeletal<br>symptoms<br>n% | 126(33.2) | 1488(26.8) | 0.007 | 130(33.1) | 2020(29.6) | 0.139 | 5(19.2) | 1115(29.9) | 0.236 |
| Systemic<br>symptoms<br>n% | 231(60.8) | 3191(57.5) | 0.210 | 235(59.8) | 3809(55.8) | 0.117 | 11(42.3) | 2110(56.6) | 0.143 |

### Association between pregnancy and symptomatic COVID-19

After multivariable logistic regression, pregnancy was associated with higher odds of symptomatic COVID-19 (OR 1.41, 95% CI 1.18-1.68, p<0.01) (Table 5). Further, pregnancy was associated with higher odds of shortness of breath (OR 1.29, 95% CI 1.02-1.63, p=0.03) (Table 5).

**Table 5:**
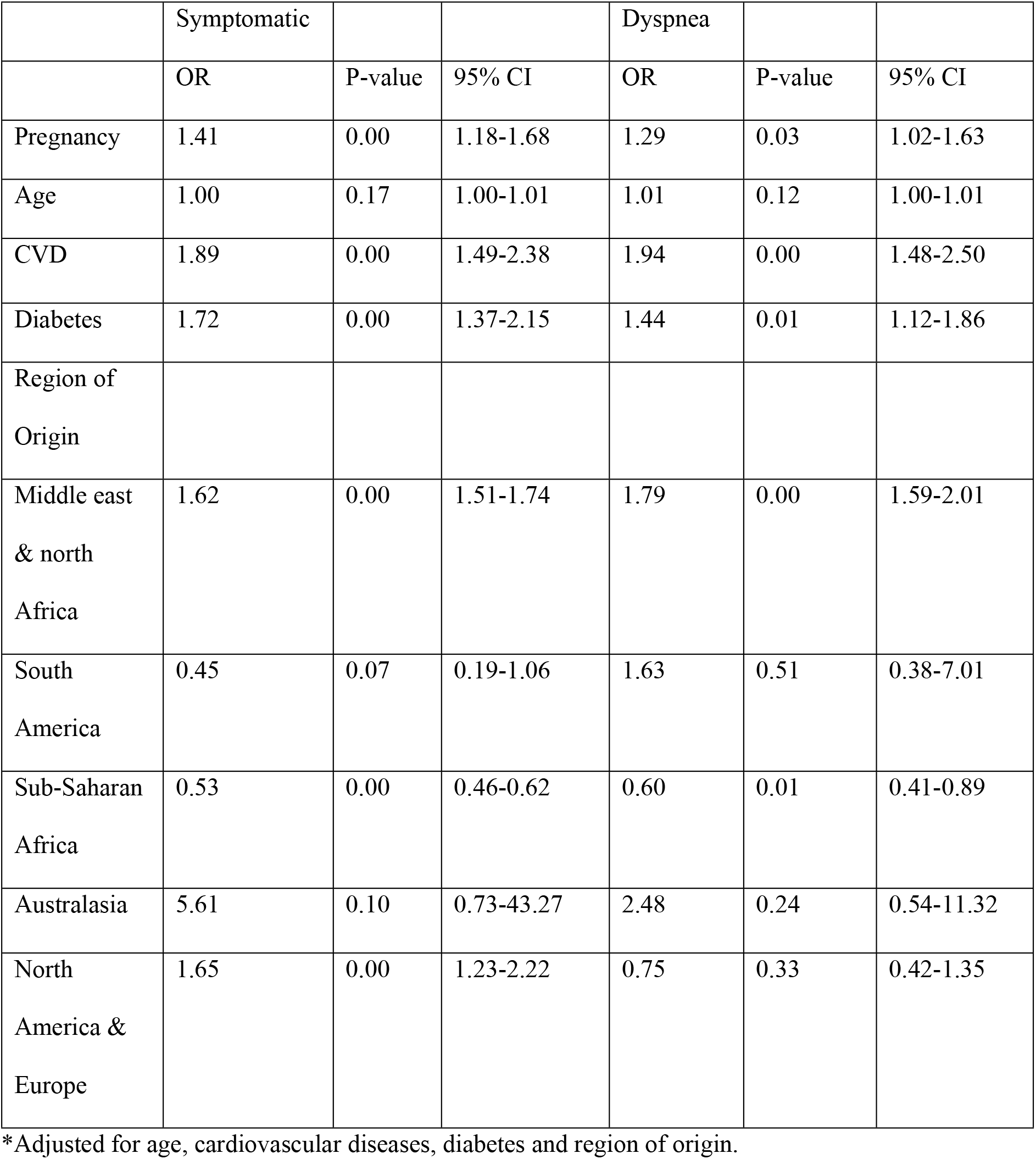
Association between pregnancy and symptomatic COVID-19 – multivariable logistic regression.

|  | Symptomatic |  |  | Dyspnea |  |  |
| --- | --- | --- | --- | --- | --- | --- |
|  | OR | P-value | 95% CI | OR | P-value | 95% CI |
| Pregnancy | 1.41 | 0.00 | 1.18-1.68 | 1.29 | 0.03 | 1.02-1.63 |
| Age | 1.00 | 0.17 | 1.00-1.01 | 1.01 | 0.12 | 1.00-1.01 |
| CVD | 1.89 | 0.00 | 1.49-2.38 | 1.94 | 0.00 | 1.48-2.50 |
| Diabetes | 1.72 | 0.00 | 1.37-2.15 | 1.44 | 0.01 | 1.12-1.86 |
| Region of Origin |  |  |  |  |  |  |
| Middle east & north Africa | 1.62 | 0.00 | 1.51-1.74 | 1.79 | 0.00 | 1.59-2.01 |
| South America | 0.45 | 0.07 | 0.19-1.06 | 1.63 | 0.51 | 0.38-7.01 |
| Sub-Saharan Africa | 0.53 | 0.00 | 0.46-0.62 | 0.60 | 0.01 | 0.41-0.89 |
| Australasia | 5.61 | 0.10 | 0.73-43.27 | 2.48 | 0.24 | 0.54-11.32 |
| North America & Europe | 1.65 | 0.00 | 1.23-2.22 | 0.75 | 0.33 | 0.42-1.35 |

\*Adjusted for age, cardiovascular diseases, diabetes and region of origin.

### Association between pregnancy and the number of COVID-19 symptoms

After multivariable Poisson regression, pregnancy was associated with a higher number of symptoms (IRR 1.03, 95% CI 0.98-1.08, p= 0.31) (Table 6).

**Table 6:**
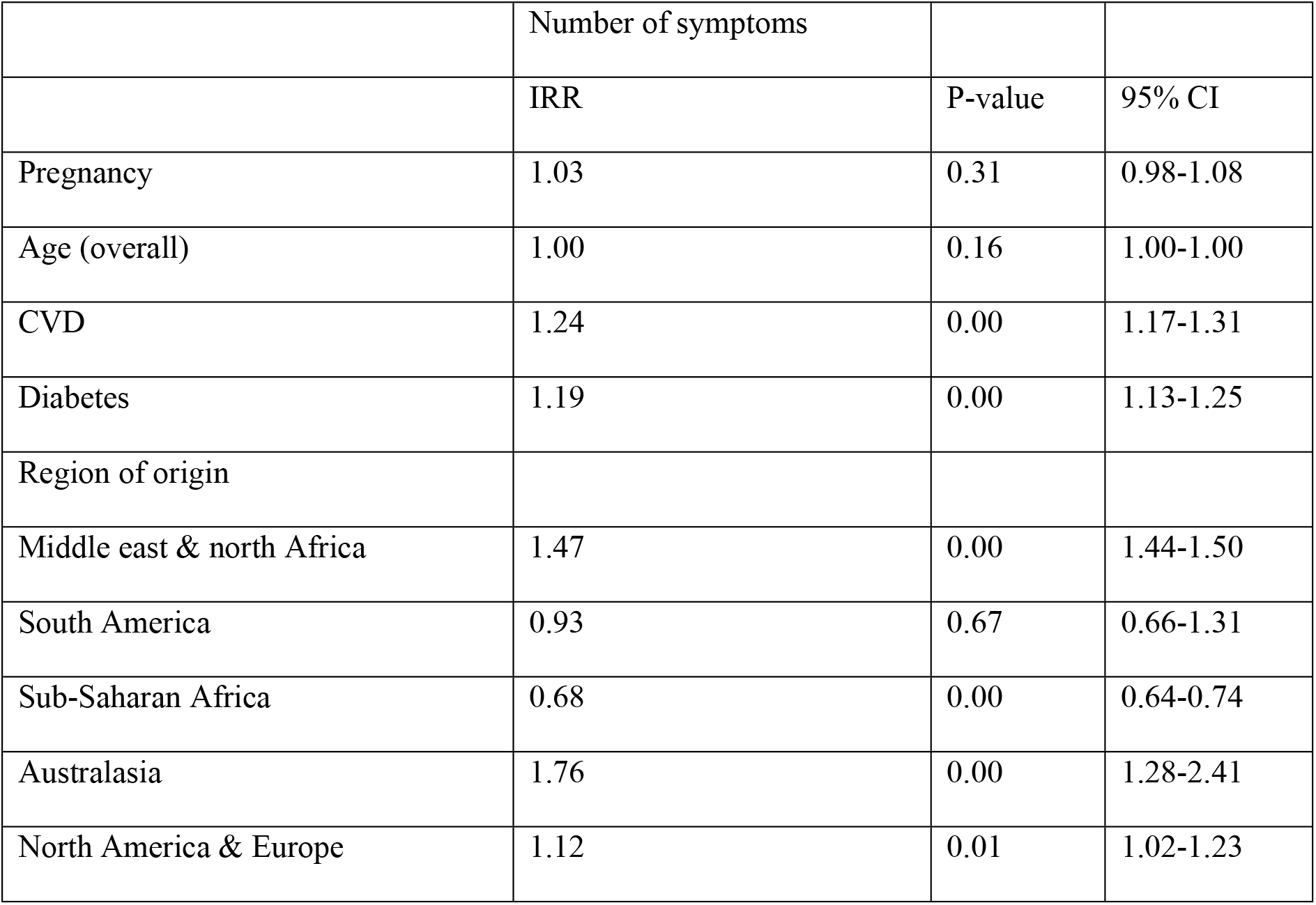
Association between the number of symptoms and pregnancy – Multivariable Poisson Regression.

## Discussion

In this population based cross sectional study, we found that, in a diverse Middle Eastern population, pregnant women were more likely to report symptomatic COVID-19 compared to non-pregnant women. Moreover, pregnant women had a higher proportion of shortness of breath, compared to women who were not pregnant. After multivariable logistic regression, pregnancy was associated with 1.41-fold higher odds of having symptomatic COVID-19 and almost 1.3-fold higher odds of reporting shortness of breath. Moreover, a multivariable poisson regression analysis showed that pregnant women were more likely to have a higher number of symptoms compared to non-pregnant women.

We found higher odds of symptomatic COVID-19 and shortness of breath in pregnant women compared to non-pregnant of the same age. Although there are some contrasting results from other studies [7, 20, 21], our findings add to a growing body of evidence that suggests that pregnant women are vulnerable to severe COVID-19 [4,5,22,23], but from a region where such data have been scarce to date. In one meta-analysis of 67 271 women, pregnant women and women in the postpartum group were twice more likely to be admitted to ICU, to have mechanical ventilation and to require extracorporeal membrane oxygen [4]. In another study from the US CDC, of 461 825 women, pregnant women three times more likely to be admitted to ICU, two-and-a-half times to receive extracorporeal membrane oxygenation, three times more likely to need mechanical ventilation and almost twice likely to die from COVID-19. In contrast, one systematic review concluded that clinical features of pregnant women with COVID-19 did not vary from those of non-pregnant adults [20]. However, this review had several limitations including a very small sample size of 114 total participants and included poor-quality studies. Similarly, two other older reviews suggested that there was no difference in COVID-19 presentation between pregnant and non-pregnant women [21], and that pregnant women were less likely to have cough, sore throat, headache and diarrhea than non-pregnant adults [7]. Notably, these older reviews included small and poorly designed studies. Taken together, our findings and those of existing studies suggest a worse course of COVID-19 in pregnancy.

Several physiological changes during pregnancy may worsen the course of COVID-19 and therefore, result in severe symptoms. A more symptomatic course of the disease could be caused by changes in the respiratory system during pregnancy that can impair the protective functions of the lungs [24]. Also, high levels of estrogen and progesterone hormones during pregnancy induce the upper part of the respiratory tract to swell, which increases the susceptibility of severe COVID-19 infection. Lastly, pregnancy is a hypercoagulable state, and this makes pregnant women more vulnerable to a severe course of COVID-19, where coagulation dysfunction is a hallmark of poor disease outcomes [25,26].

There are several limitations in our study. Firstly, data collection from participants was done through phone calls, to reduce the spread of disease. This may have introduced subjectivity in symptom assessment. However, there is no basis to believe that this subjective reporting of symptoms was differential, such that it was more likely in either pregnant or non-pregnant women. Therefore, if this subjective assessment affected the study, it would most likely have biased the odds ratios towards the null, and the expected effect if objective assessment was done, would probably show higher odds ratios. Additionally, information about the gestational age of the participants was not measured, which may have affected the symptomology of COVID-19, as immunological and physiological changes vary throughout between pregnancy trimesters. Further studies investigating the effect of COVID-19 infection on pregnancy in different trimesters are therefore needed. An important question which we were not able to answer in this research is the effect of COVID-19 on pregnancy outcomes, and future research may be needed in this setting. Despite this, our study allowed us to explore the symptomology of COVID-19 infection across a multicultural cohort, given the fact that our study included a large sample with a diversity of women of childbearing age.

## Conclusion

Our findings suggest that, in a multinational cohort in Qatar, pregnant women are more likely to have symptomatic COVID-19, especially shortness of breath, compared to women without pregnancy of the same age.

## Data Availability

Data are available upon reasonable request

## Acknowledgements

We acknowledge the Ministry of Public Health of Qatar (MoPH) for the data used in this study and the Population Medicine Department at the College of Medicine at Qatar University for methodological support.

## CRediT author contributions

**AA:** conceptualization, methodology, software, formal analysis, investigation, resources, writing – original draft, writing – review and editing, visualization.

**AH:** conceptualization, methodology, software, formal analysis, investigation, resources, data curation, writing – original draft, reviewing and editing, visualization, project administration.

**AA:** conceptualization, methodology, writing – original draft, writing – review and editing, visualization.

**EH:** conceptualization, methodology, software, writing – original draft, writing – review and editing, visualization.

**NY:** conceptualization, methodology, software, formal analysis, investigation, resources, writing – original draft, writing – review and editing, visualization.

**RA:** conceptualization, methodology, software, formal analysis, investigation, resources, writing – original draft, writing – review and editing, visualization.

**DB:** reviewing and editing

**EA:** reviewing and editing

**HA:** reviewing and editing

**MA:** reviewing and editing

**OM:** data curation, reviewing and editing

**SD:** formal analysis, reviewing and editing

**TC:** conceptualization, methodology, software, data curation, validation, formal analysis, investigation, supervision, project administration, writing, reviewing and editing,

